# Spatiotemporal Rhythmic Seizure Sources Can be Imaged by means of Biophysically Constrained Deep Neural Networks

**DOI:** 10.1101/2023.11.30.23299218

**Authors:** Rui Sun, Abbas Sohrabpour, Boney Joseph, Gregory Worrell, Bin He

## Abstract

Noninvasive dynamic brain imaging of neural oscillations provides valuable insights into both physiological and pathological brain states. Yet, challenges remain due to the ill-posed nature of the problem and high complexity of the solution space, which can be alleviated by advanced computational models. Here, we investigated the capability of a novel deep learning-based source imaging framework (DeepSIF) for imaging ictal activities from high-density electroencephalogram (EEG) recordings in drug-resistant focal epilepsy patients. The neural mass model of ictal oscillations was adopted to generate synthetic training data with spatio-temporal-spectra features similar to ictal dynamics. We rigorously validated the trained DeepSIF model using computer simulations and in a cohort of 33 drug-resistant focal epilepsy patients. The DeepSIF ictal source imaging was compared with interictal source imaging and three conventional imaging methods as benchmark comparisons. Our findings show that the trained DeepSIF model outperforms other methods in estimating the spatial and temporal information of ictal sources. It achieves a high spatial specificity of 96% and a low spatial dispersion of 3.80 ± 5.74 mm when compared to the resection region. The noninvasive source imaging results also demonstrate good coverage of seizure-onset-zone (SOZ), with an average distance of 10.89 ± 10.14 mm (from the SOZ to the reconstruction). These promising results suggest that DeepSIF has significant potential for advancing noninvasive imaging of ictal activities in patients with focal epilepsy. By providing valuable insights into the spatiotemporal dynamics of seizure activity, DeepSIF promises to help guide clinical decisions and improve treatment outcomes for epilepsy patients.

## Introduction

Neural information processing is encoded by rhythmic oscillations. Noninvasive imaging of network dynamics of such neural rhythms is of significance for elucidating the mechanism of brain function and aiding clinical management of brain disorders. Epilepsy is a common neurological disorder affecting around 70 million patients worldwide [1], one third of whom cannot be managed by medication alone. For focal drug resistant epilepsy (DRE), in which seizures are originated in a focal region of the brain, surgery to remove the epileptogenic zone (EZ) has proven to be the most effective treatment option [2]. Accurately identifying the epileptogenic tissue is of great importance to the diagnosis and treatment planning for these patients. Patients selected for surgical treatment usually undergo multi-day (sometimes even weeks) intracranial EEG (iEEG) monitoring to determine the epileptogenic tissue from invasive ictal recordings, which is a close approximation of the EZ [3]. However, iEEG sometimes is limited by its spatial coverage, as well as the risk and discomfort associated with the invasive procedure [4]. There is a clinical need for noninvasive imaging techniques with high spatiotemporal resolution, to identify and localize seizure generating tissues with high accuracy.

Electroencephalography and magnetoencephalography (E/MEG) are noninvasive techniques that can record neural activities with high temporal resolution. However, limited spatial information can be inferred from the scalp measurements because of the low signal to noise ratio (SNR), small sensor counts, and the volume conduction effects. Electrophysiological source imaging (ESI) techniques have been developed to boost the spatial resolution of E/MEG by estimating the underlying brain dynamics from E/MEG recordings [5–11]. ESI involves solving the forward problem and the inverse problem. The forward problem models the source space as a distribution of current dipoles, and its mapping relationship to the sensor space as a linear projection [12]. Since the number of scalp measurements is much smaller than the number of current dipoles, a regularization procedure is conventionally used to solve the underdetermined inverse problem. However, the performance of the ESI solution is limited by the representation power of the regularization term [13].

Deep learning (DL) - based source imaging approaches solve the ESI problem under a different framework. Instead of being formulated explicitly as a regularization term, source dynamics can be implicitly embedded in the training data and learned by the neural networks through the training process. It is, however, challenging to acquire enough simultaneously recorded iEEG and scalp data to train such a model. Computational models can serve as a powerful alternative to introduce the source dynamics into training data, if the source models are realistic enough for source imaging tasks. DL-based source imaging framework (DeepSIF) is recently proposed as a general ESI framework unitizing dynamic brain network models as the source model in the forward problem, and the deep neural networks (DNN) to solve the inverse problem [14,15]. Different source models can be selected based on source signal properties and the modalities of the measurements, to generate various types of source-sensor signal pairs as the training data. It has been successfully applied to image transient activities such as evoked potentials or interictal spike activities from EEG or MEG signals, demonstrating powerful generalization capabilities cross subjects and modalities.

On the other hand, the oscillatory activities in EEG signals are fundamental for inferring physiological or pathological information about specific brain states or disorders and have increasingly gained attention [16,17]. One example is the occurrence of strong rhythmic patterns during seizures and the origin of these patterns is a major piece of evidence used for determining the EZ. Studies have shown that ictal ESI could be more informative than interictal ESI results [18–20]. However, ictal EEG recordings are usually contaminated by large artifacts. Although independent component analysis (ICA) can remove some artifacts, ictal oscillations still have low SNR due to non-ictal brain rhythms, making the analysis of these patterns challenging [21]. To overcome these issues, several approaches have been proposed, such as transforming the data into the frequency domain [22,23], or averaging the signal at the peak of the ictal oscillations to increase the SNR of the ictal recordings [24], albeit at the expense of temporal resolution. Averaging the EEG over time, while effective in improving SNR, can reduce the spatial specificity of the ESI results. Thus, it is important to develop a robust spatiotemporal imaging method that can model the ictal neural oscillations when analyzing the ictal EEG data.

As a general ESI framework, DeepSIF has been shown to provide excellent spatiotemporal ESI results on non-oscillatory activities at low SNR. In this work, we expanded the capability of DeepSIF to imaging oscillatory activities, specifically, the ictal oscillations. The overall study design is illustrated in Fig. 1. Ictal spatial and temporal source models were developed to generate synthetic training data for the DNN. The trained network was then utilized to image and localize seizure generating tissues from scalp recorded high density ictal EEG in 33 focal DRE patients. To evaluate the performance of DeepSIF in imaging ictal activities, the resulting images were compared to iEEG defined seizure onset zones (SOZ) and/or resection volumes. In addition, the DeepSIF based ictal source imaging is rigorously evaluated against interictal source imaging, as well as three conventional source imaging methods. The present study found that DeepSIF could successfully image ictal activities with a high degree of temporal correlation with the scalp recordings and high spatial precision that concorded with the clinical ground truth. These results suggest that DeepSIF has great potential in advancing the noninvasive imaging of ictal activities in patients with focal epilepsy, which could provide valuable insights to guide clinical decisions and improve treatment outcomes.

**Figure 1.**
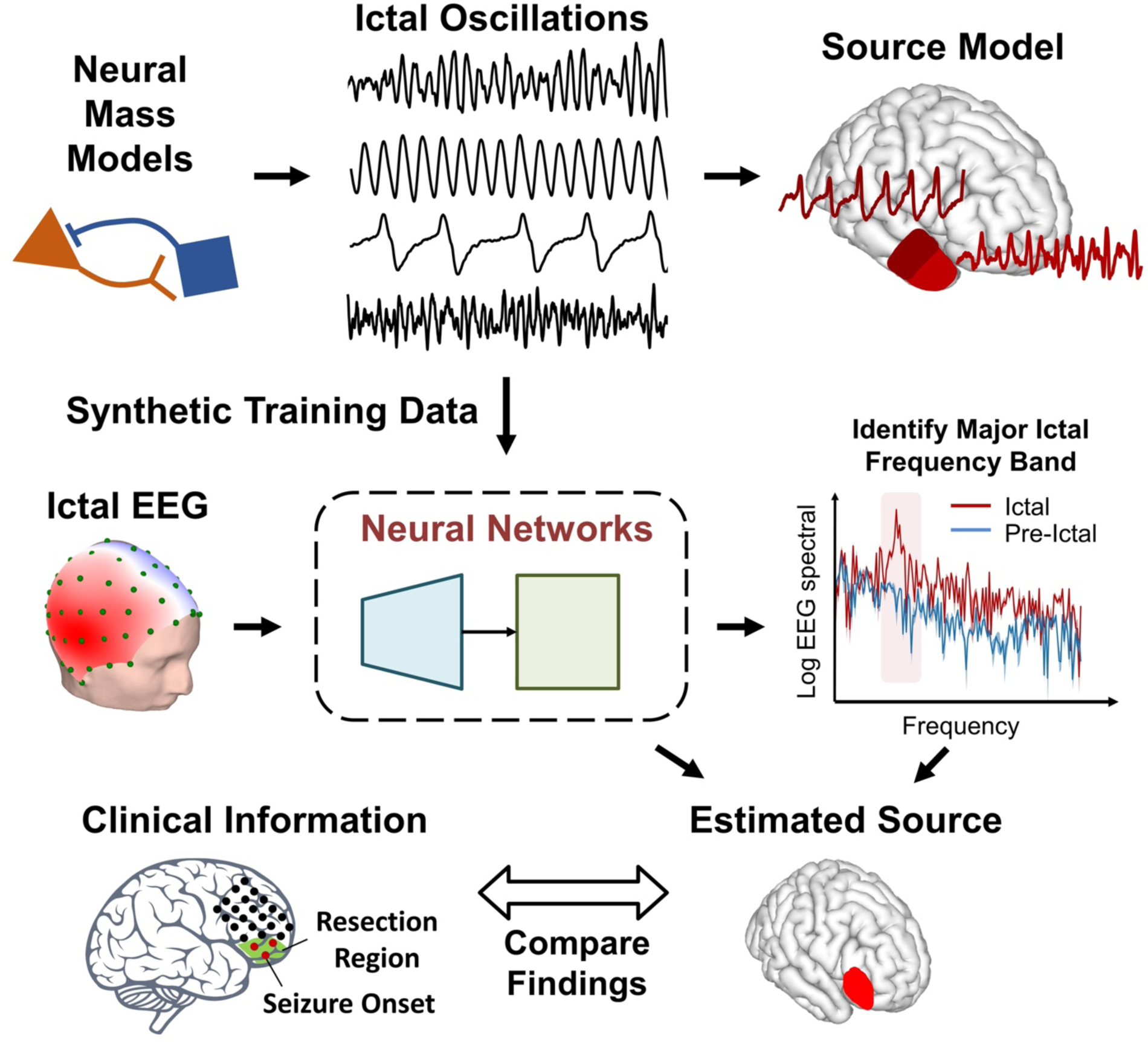
Schematic diagram of study design. Brain activities are modeled by the spatiotemporal source model, consisting of interconnected neural mass models. Ictal oscillations are generated and projected to the scalp to train a deep neural network. The trained model can be directly used to estimate seizure dynamics at the major ictal frequency band.

## Results

### Ictal Oscillation Simulations

The modified Jansen-Rit model [25], a neural mass model (NMM), was adopted as the temporal model for the ictal signal. Fig. 2a shows the typical waveforms in each of the 6 signal groups: normal activity, sporadic spikes, sustained discharge of spikes, rhythmic activity, low voltage rapid activity and quasi-sinusoidal activity. A typical example of the NMM simulation result is shown in Fig. 2b. The color represents the signal type transition when varying the time constant *a* and *b*. With these sets of NMM parameters, NMM can generate five types of signals typical in ictal oscillations with different temporal and spectra features. Fig. 2c shows the waveforms generated by the corresponding *a*, *b* parameters in Fig. 2b. Although the signals share similar temporal features within each signal type/category, their major frequency gradually changes with different time constant values, providing more variations for the training dataset.

**Figure 2.**
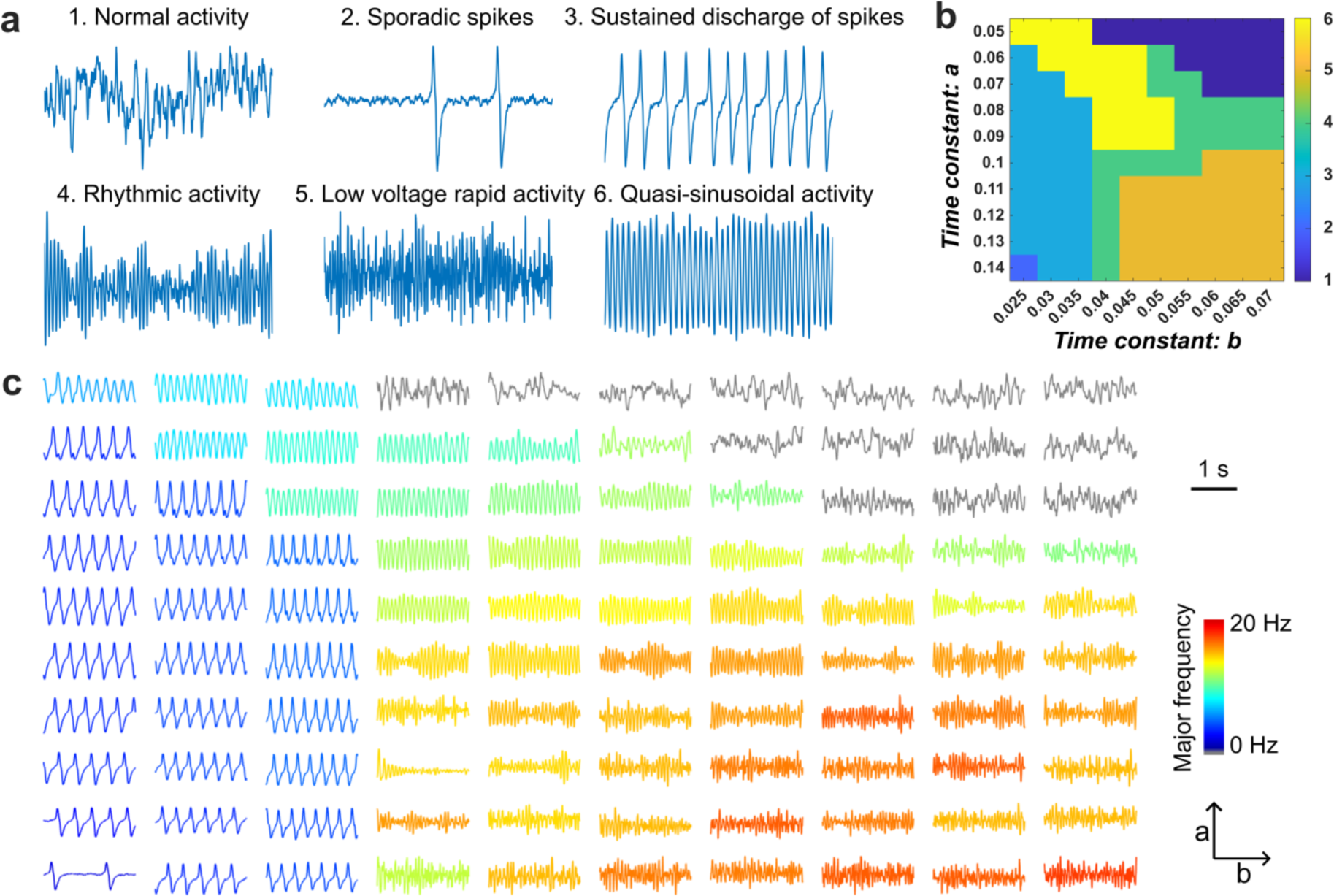
Ictal oscillation patterns generated by the modified Jansen-Rit Model. **a**, 5 seconds of typical waveforms for each signal type. **b**, Signal type classification example when varying the *a*, *b* values. The signal type labels 1-6 correspond to the event labels in a. **c**, 2 seconds of waveform examples when varying *a*, *b* values. The color denotes the major frequency of the ictal oscillations defined as the frequency with maximum power. Grey color denotes the normal activity, and the major frequency is not calculated.

### DeepSIF Performance in Computer Simulations

A DeepSIF model with skip connections and recurrent layers was trained with synthetic ictal data, comprising of source-sensor paired signal activities generated by the NMM. Then, the ability for DeepSIF to detect temporal dynamics variations in time and in space was evaluated on two test datasets and is shown in Fig. 3. High spatial specificity and sensitivity can be achieved for both test datasets. When the oscillation pattern varies over time, DeepSIF can identify the change and provide the correct temporal estimation with high linear correlation with the simulated signal (0.98±0.04). Note that in the training data, ictal type remains constant over time for one training sample. However, due to the presence of various ictal types in the training dataset through different samples, the DNN still has the capability to accurately image the source even when the oscillation pattern varies. It is more challenging when two different dynamics are presented in one patch. As illustrated in the example in Fig. 3b, a single patch contains two sinusoidal oscillations with a phase difference, and DeepSIF can provide distinct temporal estimates for each source. At around 200 ms, the source signal phases are reversed, resulting in less accurate estimations from DeepSIF due to signal cancellation in the sensor space. However, the recurrent structure allows DeepSIF to utilize information from previous time points in order to provide more reliable estimations for ambiguous time points. The temporal correlation of the simulated and estimated signal remains high (0.92±0.07) for this test dataset, despite the temporal inferences inside the source patch. This highlights the model’s ability to adapt and maintain accuracy even in challenging situations. The aggregated results for the two test datasets at different SNR are described in Fig. 3c, demonstrating consistent performance across all SNR levels. The simulation study demonstrated that DeepSIF can deliver accurate and robust source estimates for sources exhibiting a wide range of spatiotemporal patterns, emphasizing its reliability and adaptability in varying conditions.

**Figure 3.**
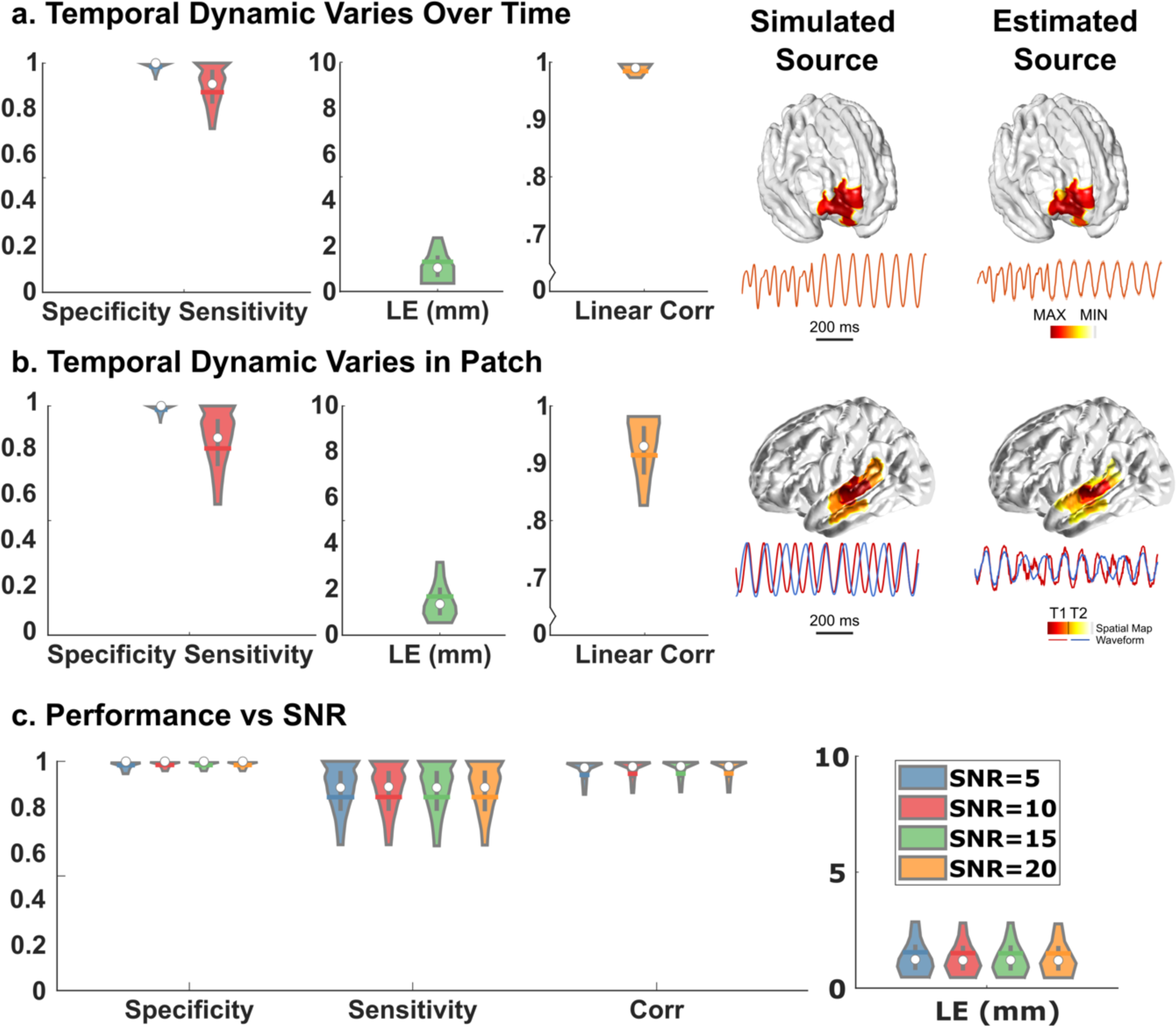
Model performance in simulations. **a**, Results on test dataset with temporal dynamics varying in time (n = 23,856). Left: Simulation results. Spatial specificity, spatial sensitivity, localization error, and temporal correlation. The distributions are demarcated within the 10th to 90th percentile. The gray bars span the 25th to 75th percentile, the white circle is the median and the colored horizontal bar is the mean of the distribution. Right: Imaging examples. Source locations and waveforms (1 second) of the simulated and reconstructed sources are plotted. **b**, Results on test dataset with temporal dynamics varying in space. Left: Simulation results. Spatial specificity, spatial sensitivity, localization error, and temporal correlation. Right: Imaging examples. Two dynamics are presented in one patch denoted by red and yellow. The reconstructed spatial maps for the two dynamics (T1 and T2) are denoted by the colorbar and the temporal waveform are plotted in the corresponding color as the simulated source. **c**, Imaging performance for the two datasets at different SNR.

### Patient Data Analysis

The DeepSIF performance for imaging real ictal signals was evaluated in a cohort of 33 focal DRE patients. Fig. 4b presents examples of ictal imaging, showcasing the excellent performance of DeepSIF when compared to surgical resection outcomes and iEEG-defined SOZ. A high spatial specificity is obtained (0.96 ± 0.90), indicating that the noninvasive DeepSIF source imaging results have minimum spurious activities extending outside the epileptogenic region, as also evident by the low spatial dispersion value (3.80 ± 5.74 mm). The average distance from the SOZ electrode to the reconstruction area is 10.89 ± 10.14 mm for all patients and 8.03 ± 9.01 mm for seizure-free patients. Furthermore, the reconstructed waveform exhibits a high temporal correlation (0.81 ± 0.14) with the EEG signals, demonstrating DeepSIF’s capability in reconstructing the temporal dynamics from EEG traces.

**Figure 4.**
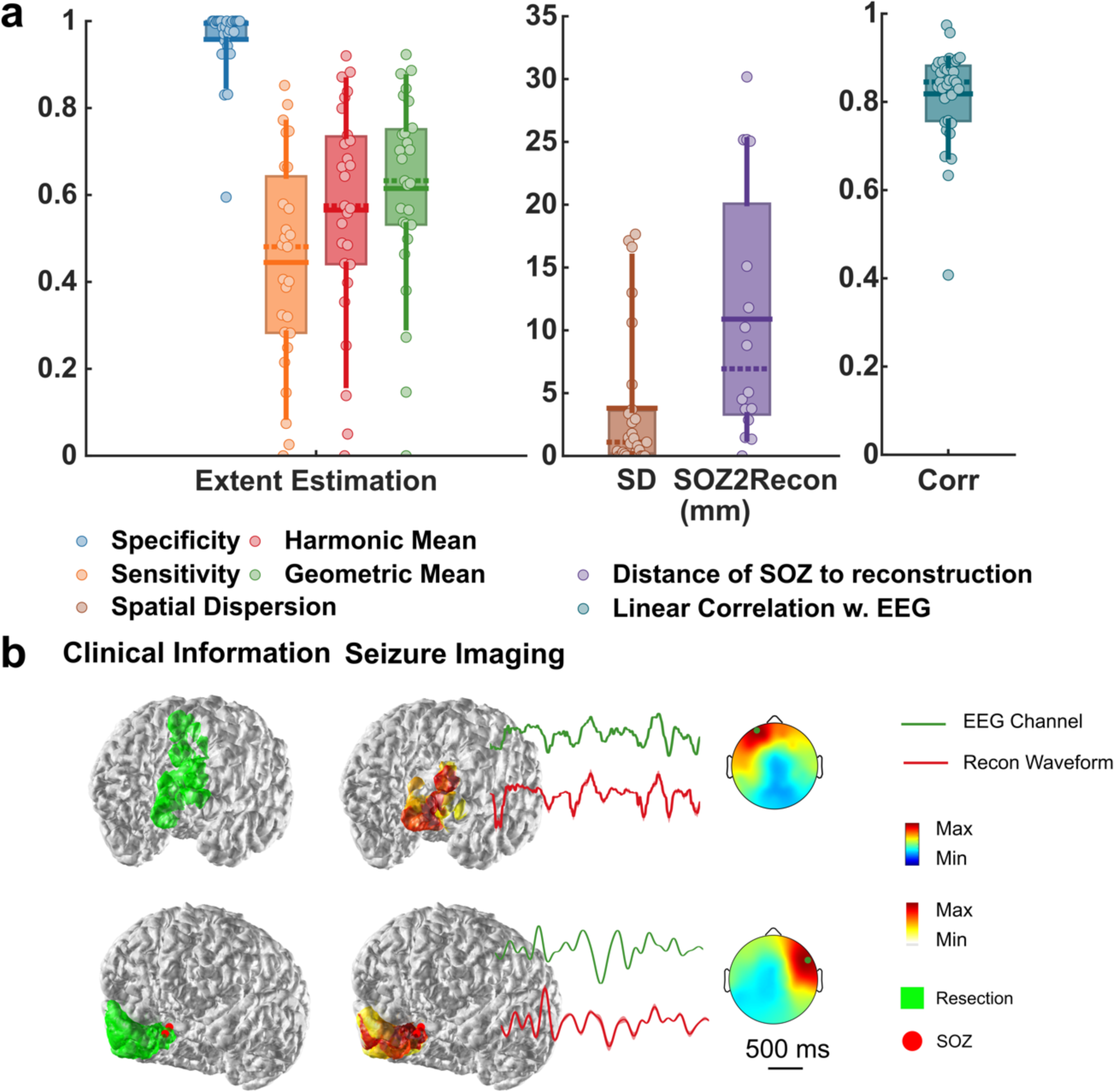
Clinical validation in drug-resistant epilepsy patients in comparison to surgical resection and iEEG-defined SOZ. **a**, Quantitative ictal-imaging results. The horizontal solid line shows the mean, the dashed line shows the median, the boxes span the 25th to 75th percentile of the data, the vertical bars span the 10th to 90th percentile of the data, and each circle represents individual patients. **b,** Examples of ictal-imaging results along with the surgical resection outcome and iEEG defined SOZ. The green channel is the channel with the maximum energy and its traces are plotted in green. The topographical maps are plotted at the maximum time point of the segment.

When comparing the results of ictal and spike imaging, statistically significant differences can be observed for all metrics except spatial dispersion. The spatial dispersion for spike imaging exhibits a bimodal distribution and can be divided into seizure-free (SZ-free) and non-seizure-free (non-SZ-free) groups. The spatial dispersion value is significantly lower for ictal imaging compared to spike imaging in the non-SZ-free group. This difference in significance can also be observed in the non-SZ-free group for spatial sensitivity and specificity. In general, the primary distinction between ictal imaging and spike imaging lies in the non-SZ-free group. This is predominantly due to the fact that many non-SZ-free patients experience multiple types of spikes, several of which are contralateral to the clinical ground truth. Even when the source can be lateralized before spike analysis, it remains challenging to differentiate between different spike groups, as demonstrated in the example shown in Fig. 5e. On the other hand, seizure onset locations tend to be more consistent, with fewer contralateral or discordant seizures. This consistency highlights the value of ictal imaging in providing a more accurate representation of seizure activity, particularly for non-SZ-free patients. Fig. 6 shows the comparison of DeepSIF with other benchmark ESI methods: sLORETA, FDI, and LCMV. DeepSIF demonstrated superior imaging performance, achieving a SOZ LE of 16.94 ± 9.08 mm, which is significantly better when compared to the benchmark methods. While LCMV and sLORETA can provide high sensitivity, their low specificity hampers their ability to accurately identify true epileptic regions. DeepSIF offers a balanced sensitivity and specificity value, allowing for a more accurate representation of the source extent without being excessively focal or overly diffused. DeepSIF offers a balanced sensitivity and specificity value, allowing for a more accurate representation of the source extent without being excessively focal or overly diffused. The geometric mean of the sensitivity and specificity can reflect the overall performance for estimating the resection regions, and it was 0.62 ± 0.22 (DeepSIF), 0.60 ± 0.15 (sLORETA), 0.54 ± 0.16 (FDI), and 0.47±0.17 (LCMV). Precision and recall are also important metrics to evaluate the overlap between two areas. The sensitivity is the same as the recall. The precision value compared to the resection area and the F1-score (the harmonic mean of the precision and recall) are shown in Supplementary Fig. S3, demonstrating that DeepSIF outperformed all other benchmark algorithms in terms of source extent imaging.

**Figure 5.**
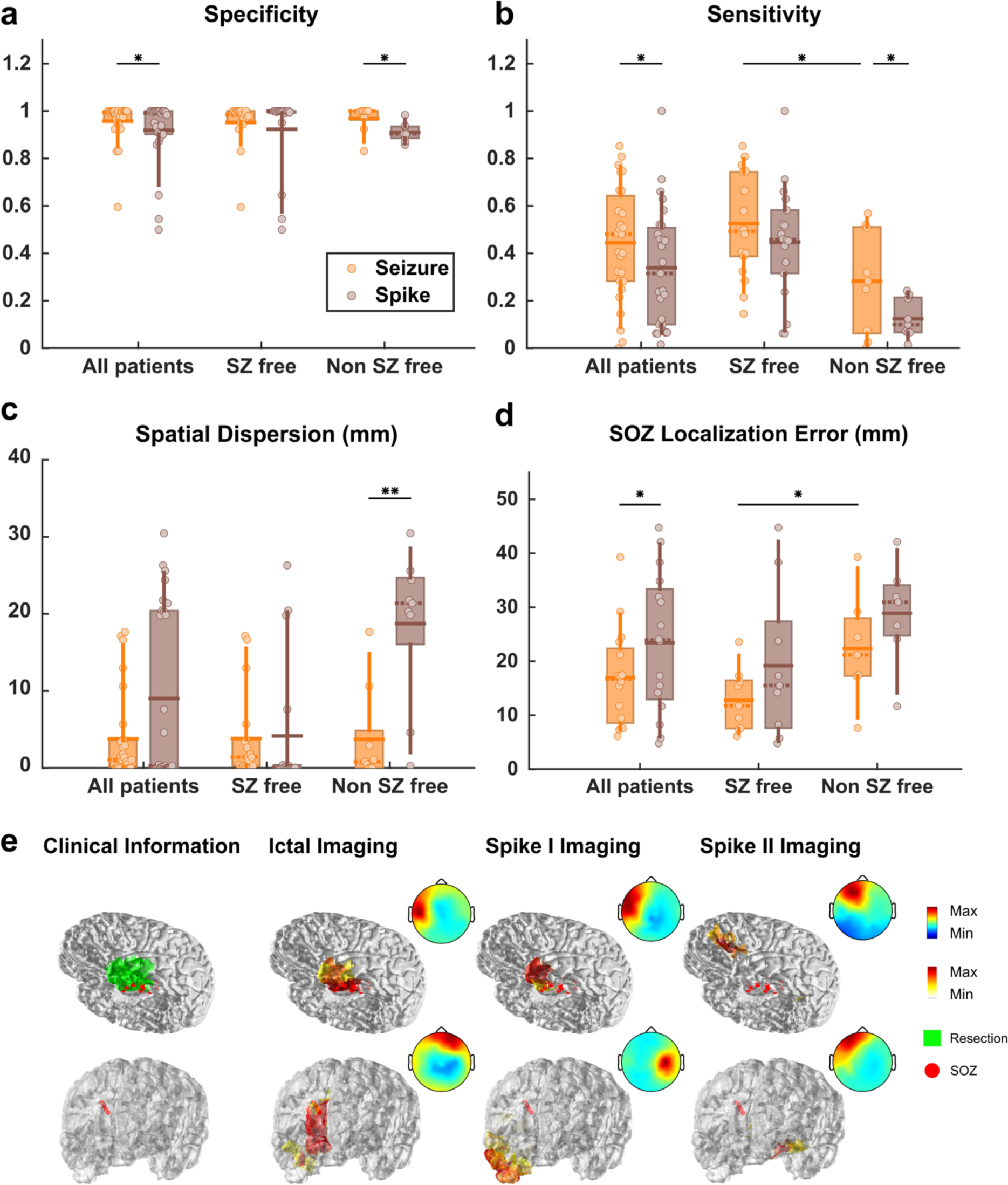
Comparing ictal imaging with spike imaging results. **a**, Spatial specificity. **b**, Spatial sensitivity. **c**, Spatial dispersion (mm). **d**, SOZ localization error (mm). Paired one-sided Wilcoxon signed rank test was used with statistical significance cutoffs of (*P<0.05, **P<0.01). **e,** Examples of ictal and spike imaging results.

**Figure 6.**
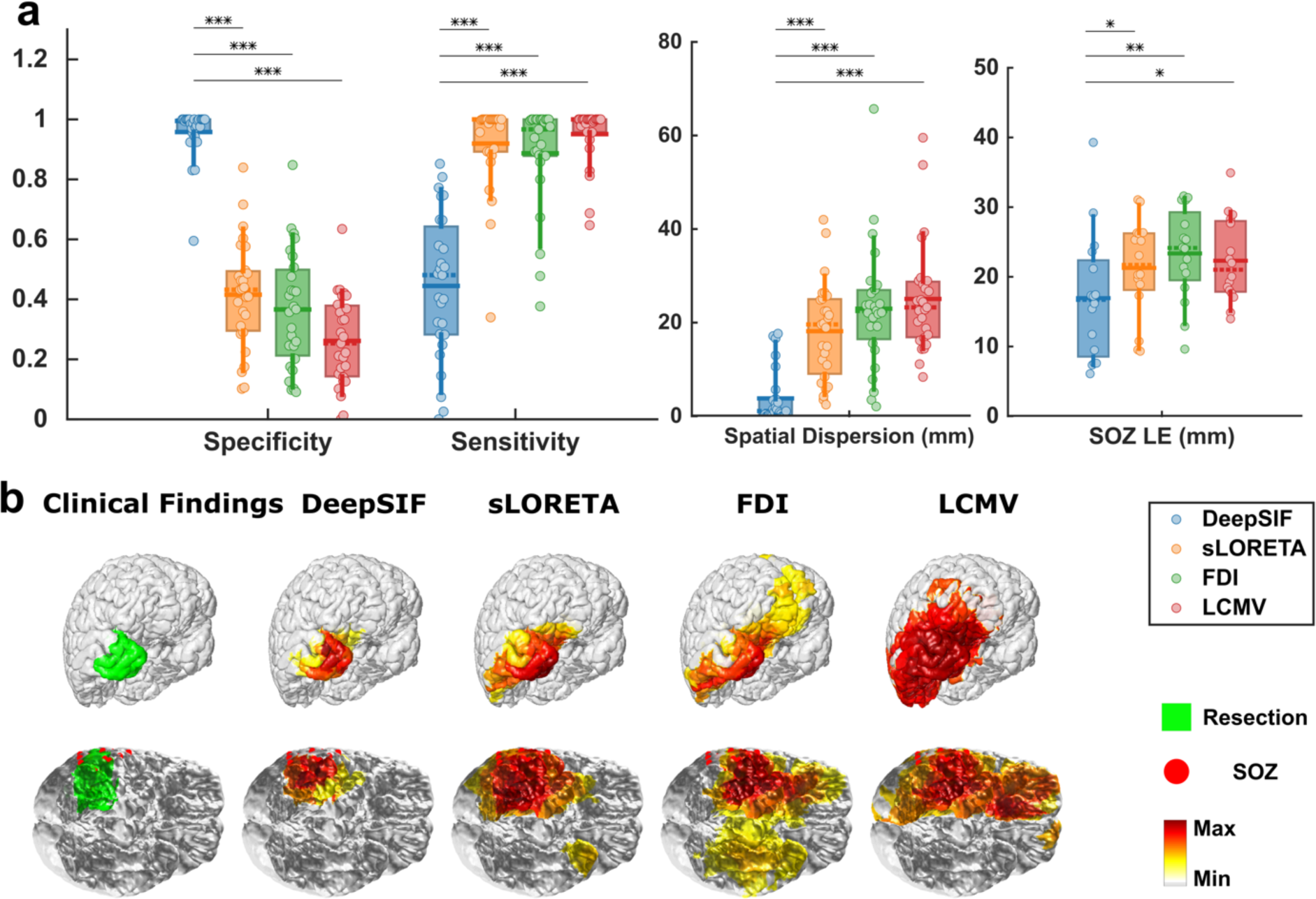
Ictal imaging results for DeepSIF, sLORETA, FDI, and LCMV. **a,** Spatial specificity; Spatial sensitivity; Spatial dispersion (mm); SOZ localization error (LE) (mm). Paired one-sided Wilcoxon signed rank test was used with statistical significance cutoffs of (*P<0.05, **P<0.01, ***P<0.001). **b,** Examples of spike-imaging results along with the surgical resection outcome and iEEG defined SOZ.

## Discussion

We have developed a novel EEG source imaging approach by means of biophysically constraint deep neural networks, to robustly and accurately localize and image spatiotemporal distribution of seizure sources from EEG ictal recordings. The method has been rigorously validated in a cohort of 33 drug resistant focal epilepsy patients by comparing noninvasive source imaging results with clinical ground truth based on intracranial EEG defined seizure onset zone and successful surgical resection outcome. We also demonstrate the superior performance of our DL-based ictal source imaging approach with well-established conventional source imaging methods, and with interictal spike source imaging.

DL-based ESI methods have shown great promise over the last few years. These methods are capable of implicitly learn the source distributions through data instead of explicitly formulating the regularization terms to constrain the solution space, which means more complex source models can be incorporated into the solution to achieve a more accurate and robust source estimate. There have been several recent attempts to image brain activities using deep neural networks [26–31]. They have shown excellent performance in computer simulations, demonstrating the power of DL-based ESI methods. DeepSIF as a DL-based ESI method, has proven to be effective for imaging transit activities such as interictal spikes or evoked potentials in a large group of subjects [14,15]. It is a modular framework consisting of a forward source model, using neural mass models, to generate realistic synthetic training data, and an inverse neural network model to perform the ESI task based on the information in the training data. The two components are closely connected though the training data in a sense that enough high-quality training data needs to be fed into the neural network for an optimal training result. However, they are also independent as the detailed implementations and assumptions in the source model are implicitly embedded into the training data and the complexity of the source model will not affect the optimization process for the inverse module. Thus, researchers can adopt different forward source models and inverse network structure for DeepSIF based on their tasks to achieve the optimal results.

Current development of DL-based ESI methods focuses on the spatial localization or imaging the transient spiking activities. Yet, noninvasive imaging of the spatiotemporal rhythmic signal has gained significant attention in recent years, as it offers valuable insights into the physiological and pathological brain states [16,17]. One important application is the recording and analysis of ictal activities during the presurgical evaluation for DRE patients in order to determine the EZ. Currently, invasive iEEG monitoring serves as the clinical gold standard for identifying the epileptogenic tissue [32], but the success rate of iEEG guided surgical treatment is limited when iEEG electrodes fail to cover the EZ. An accurate noninvasive ictal imaging technique could offer a clear hypothesis regarding the EZ area, which would subsequently facilitate iEEG implantation and may ultimately lead to the complete noninvasive source localization of seizure generating tissues, potentially leading to improved patient outcomes. Given the potential benefits in epilepsy treatment, extending DeepSIF to imaging rhythmic activities is of great importance, particularly for imaging ictal activities. As it has demonstrated promise in localizing the sources of brain activity with high accuracy and robustness, by developing DeepSIF for ictal activities, it is possible to create a more comprehensive and reliable methodology for identifying EZ noninvasively.

To generate the training data for the ictal DeepSIF model, a modified Jansen-Rit model was adopted for generating training data with various spatio-temporal-spectra features. Multiple computational models have been proposed to describe and analyze the seizure dynamics [33]. Concepts of increased tissue excitability, impaired dendritic inhibition, coupling interactions have been incorporated into models at various scales to provide insights regarding the biophysical mechanisms of seizure initiation, propagation and termination at micro-to-macro scales [25,34–39]. Developing a seizure model that can fully describe the seizure process is an activate research area. Certain simplifications were made when selecting and constructing the source model for DeepSIF, with the primary focus being on reproducing realistic phenomena observed in the recorded signals, rather than elaborating on the underlying mechanisms. This approach prioritizes the practical application and performance of DeepSIF in real-world situations. First, the neural mass model (NMM) needs to be able to generate a wide range of ictal dynamics. Models have been proposed to model and explain certain types of ictal oscillations [35,37]. The modified Jansen-Rit model have demonstrated its ability to model different types of ictal oscillations [25], with a wide range of parameter sets for each type of signal. A large number of parameter sets were explored and included in the training data, making it a suitable model to build a training dataset with high varieties. Second, the parameters were manually modified for the transition between ictal signal types. Some models can autonomously transit between dynamical states by setting the NMM system close to the bifurcation and providing random fluctuations as the input [34,37]. However, this limits the possible parameter selections, thus the possible waveform dynamics in the training data. As a trade-off, the transition between states were modeled by manually changing the NMM parameters to generate different ictal signal types. Similarly, the spatial dynamic variations are also modeled through manually changing the parameters. Only a short time segment of seizure was considered in the source model. As a seizure could last from a couple seconds to couple minutes, it is challenging to include all the possible temporal variation patterns and spatial propagation patterns in the training data through the whole course. We simplified the problem and assumed the ictal source is piece-wise stationary, which means in a short segment of time, the source is confined in a focal region. Then, the size and shape of the seizure source can be properly defined by a patch, and the spatial dynamic variations can be introduced by manually modifying the signal types in the center segments and neighboring segments of the source patch (Fig. 2b). These assumptions and simplifications provide a valid trade-off between the realism and the implementation feasibility of the source model, and it is suitable for the source imaging tasks.

Generating realistic datasets has been one of the bottlenecks for the advancement of DL-based ESI methods. The development of the spatiotemporal ictal source model provides a training dataset with enough spatio-temporal-spectra features, which is a critical step to provide an accurate ictal imaging result and demonstrated the capability of DeepSIF for seizure source imaging. The DeepSIF model was trained on a generic head model, and it was rigorously validated in computer simulations and in a cohort of 33 focal DRE patients. The trained DeepSIF model demonstrates superior robustness and generalizability on various test conditions, and it can be successfully applied to different patients with certain spatiotemporal robustness, which provides a high level of efficiency as single trained model can be directly applied at multiple test conditions. The model can accurately estimate the spatial and temporal information of the ictal sources, providing a high temporal correlation value of 0.95 ± 0.07 in computer simulations, and of 0.81 ± 0.14 in patient data analysis. The model can also provide high spatial specificity (98% in simulations and 96% in patient data analysis) with decent spatial sensitivity (84% in simulations and 44% in patient analysis), which means the model can provide accurate localization without having nuisance spurious activities extended outside the resection region. Multiple factors can influence the resection volume in practice, including imaging and functional testing findings, patient’s treatment goals, encountered individual anatomical reality during an operation, neurosurgeon’s assessment of risks, the resection region is not necessary the ground truth for the ictal activities [40,41], however, it is still a valuable benchmark to validate our imaging results. It is challenging to achieve both high sensitivity and high specificity, as there is a tradeoff between providing sparse or diffused solutions. The well-established conventional ESI methods are known to have high sensitivity but low specificity. DeepSIF has a low sensitivity compared to some diffused conventional methods (like sLORETA and LCMV), however, DeepSIF has a significantly higher specificity, leading to a higher geometric mean value compared to benchmark methods. On the other hand, the ictal imaging results from DeepSIF had a good overlap with the iEEG SOZ electrodes. As the SOZ LE is defined as average of distance of each SOZ to the closest reconstruction and the distance of each reconstructed region to the closest SOZ, a solution needs to be close to the SOZ electrode groups and be neither too diffused nor too focal to achieve a low LE value. DeepSIF reached a LE of 16.94 ± 9.08 mm for all patients and 12.75 ± 5.80 mm for seizure free patients, which are statistically significantly smaller than the benchmark methods.

Our results also show that ictal imaging can more reliably estimate epileptogenic tissue compared to spike imaging. Several studies have demonstrated the advantage of ictal imaging over spike imaging [18,19,42]. By comparing the DeepSIF ictal and spike imaging results, we found that the ictal imaging is statistically significantly more accurate than the spike imaging in terms of sensitivity, specificity and localization error based on iEEG defined SOZ. When separating the patient groups into seizure free and non-seizure free groups, it can be observed that the performance difference is mainly caused by the multiple interictal spike types, while seizure sources are usually consistent and ipsilateral to the clinical ground truth. Previous studies have also observed that interictal spike clusters could be discordant with iEEG findings [43,44], and spike imaging would fail to accurately identify the EZ in these cases. Ictal imaging results become more critical in this case to resolve the inconsistency in spike imaging. Our ictal imaging approach can provide valuable information during the surgical evaluation process regarding the location of the SOZ and EZ, by providing an accurate ictal imaging result with high spatial specificity.

In sum, we have demonstrated that DeepSIF can provide robust extent, location, and temporal dynamics estimation for imaging ictal oscillations from scalp EEG in numerical experiments and real data analysis in 33 drug-resistant focal epilepsy patients. As a DL-based ESI method, it has the advantage of fast inference and no parameter tuning during the evaluation phase, while providing accurate and robust imaging results. The model’s adaptability and reliability in handling diverse ictal patterns make it a promising tool for advancing noninvasive imaging of ictal activities in patients with epilepsy.

## Methods

### Ictal Oscillation Simulations Using Modified Jansen-Rit Model

The neural mass model (NMM) is a “mean-field” computational model that describes the collective dynamics and interactions among groups of neurons and has been widely used as the source models for iEEG/EEG/MEG measurements [45,46]. Different types of NMMs have been proposed to model epilepsy-related activities [34,37,47]. Jansen-Rit and its modified version are among the most popular NMMs with physiologically meaningful parameters. The Jansen-Rit model contains three neural subpopulations, the primary neurons, the excitatory, and the inhibitory feedback interneurons [48]. Fast inhibitory interneurons are added in the modified Jansen-Rit model to model different types of ictal oscillations [25]. Each subpopulation is characterized by a dynamic impulse response function *h*(*t*) that transforms the pre-synaptic information (the average action potential firing rate) to the post-synaptic information (the mean membrane potential), and a static nonlinear activation function that transforms the mean membrane potential to an average firing rate. It has been shown that by modifying the synaptic gain parameters in the impulse response function, 6 types of signals can be simulated with the modified Jansen-Rit model: Normal activity (Type 1), sporadic spikes (Type 2), sustained discharge of spikes (Type 3), rhythmic activity (Type 4), low voltage rapid activity (Type 5) and quasi-sinusoidal activity (Type 6) [25,38]. Detailed NMM structures can be found in Supplementary Fig. 1.

We explored the impact of the gain parameter *A, B, G* and time delay constant *a, b, g* in the impulse response function on the signal dynamics. To further reduce the parameter search space, simulations were first performed for different *B* and *G* parameters. Generated signals were then classified into 6 groups based on line-length, major frequency, and baseline voltage as proposed in [38]. *B, G* pairs generating normal activities were removed from further simulations. Then for each pair of valid *B, G*, a grid search of *a, b, g* was performed. Five seconds of signal were generated with step size of 0.5 second for each parameter set in theVirtualBrain [46]. A total of around 210,000 sets of simulations were performed. Note that this is not an exhaustive exploration of the full parameter space of the modified Jansen-Rit model. The goal is to identify enough variations of the NMM parameter sets for the training data generation. Signal types 2-6 were possible ictal dynamics and were included in the training data. To ensure a uniformed representation of different types of ictal signals, parameter sets for each signal type were resampled to 10,000. A total of 50,000 parameter sets were identified. 40,000 sets were used as the candidate parameter sets to generate the ictal training data, and 10, 000 sets were used for the ictal testing data.

### Model Training and Evaluation

After identifying the parameter sets for a single NMM to generate the ictal oscillations, they were combined with the spatial-temporal source model to simulate the EEG signals during the ictal periods. A template magnetic resonance imaging (MRI) (fsaverage5) [49] was used to generate the head model and its cortical surface was segmented into 994 regions with each region modeled by one modified Jansen-Rit model. The spatial model is the region growing method described in [15], where the center segment was chosen randomly to determine the source location, and the source patch was created by randomly grouping the neighboring segments with the center segment. Each source patch consisted of two types of spatiotemporal dynamics. In the first type, the entire patch shared the same temporal waveform. In the second type, the source patch was separated into the center segment group and the neighboring segment group (Fig. 1, top right). Two groups had different temporal waveforms to simulate the phase differences or change of dynamics after the signal propagates to neighboring regions. The size of the center segment group was randomly selected, and the segments included were chosen using the region-growing method. The remaining cortical segments in the source patch comprised the neighboring segment group. The NMM parameter for the source patch (entire patch, or the center/neighboring segment groups) was selected randomly from the parameter sets obtained from the previous step. Thus, source patches with different sizes, shapes, locations, and temporal dynamics were generated. The 76-channel electrode layout [50] based on a 10–10 montage was used as the EEG electrode configuration by projecting the template EEG cap onto the scalp surface of fsaverage5. The lead-field matrix was calculated using the 3-shell boundary element method (BEM) model with openMEEG [51] in Brainstorm with default settings [52].

The network consisted of a spatial module to pre-filter the EEG signal and a temporal module to model the temporal dynamics. Detailed designs can be found in [15]. Both the source and sensor space signals were scaled by their maximum absolute value to have a maximum or minimum of 1 or -1. During training, the loss function was the mean square error loss (MSE) between the model output and the ground truth source activity. The whole network was implemented in PyTorch and trained on one NVIDIA Tesla V100 GPU [54].

The source patches in the test dataset were separately generated following the same protocol as the training data with different NMM parameters. Two test datasets were created with different temporal dynamics. In the first test dataset, the entire patch shared the same temporal waveform, but the ictal signal types changed within one test sample. In each test sample, the switch in the waveform dynamics, happened at a random time point. In the second test dataset, similar to the training set, there were different temporal waveforms in one source patch. Each dataset contains single source data with 23,856 samples at 5-20 dB SNR levels. Samples in these two datasets were used as the input for the trained model, and the examples and results are shown in Fig. 3.

The Otsu’s thresholding technique [55] was used to identify the boundary of the imaged source distribution. Modified spatial sensitivity and specificity [14,56] were used to evaluate the extent estimation accuracy. The localization error (LE) is defined as the average of the distance from the estimated source to the ground truth and the distance from the ground truth to the estimated source [57]. The correlation value is defined as the maximum Pearson correlation between reconstructed and simulated waveforms. One test sample consists of multiple cortical regions and the LE and correlation for one test sample is the mean values for all regions in the reconstructed source.

### Ethics Statement

Our clinical study including data collection and data analysis was approved by and performed in accordance with the regulations of the Institutional Review Boards (IRB) of Carnegie Mellon University and Mayo Clinic, Rochester. Patients gave their informed consent to participate in this study.

### Clinical Data Analysis

Thirty-three focal drug-resistant epilepsy patients (20 females; ages 32 ± 14 y) were included in this study (Supplementary Table S1). All patients underwent EEG monitoring and a resective surgery at Mayo Clinic, Rochester. High-density EEG electrodes (76 in total) were glued individually following a 10–10 montage with the reference electrode at CPz. The EEG signals were recorded using the Xltek EEG amplifier (Natus Medical Incorporated, CA, USA) at 500 Hz sampling rate. Twenty-nine patients underwent iEEG implantation. Sixteen of them had the computed tomography (CT) images to localize the electrode locations, and 27 of them had the post-operative MRI to identify the resection region. The co-registrations of the pre- and post-CT and MRI images were performed in Curry (Compumedics, NC, USA). The outcome of the surgical intervention was scored based on the ILAE system by the physicians during the follow-up period (17 ± 7 months). Twenty-one patients were seizure-free (ILAE 1-2) after the resection.

The pre-processing follows the steps described in [15,19]. The onset and offset times for each ictal activity were identified, and EEG signals between 10 seconds prior to the seizure onset, and 10 seconds after the seizure offset time were extracted for preprocessing. If the seizures lasted for more than 2 mins, only the first two minutes were extracted. The initial filtering of 0.5 – 40 Hz was first performed. Independent component analysis were performed in EEGLAB [58] on the seizure segment to remove the eye and muscle artifacts. Other components were combined to produce the clean EEG recordings. The first three seconds [59,60] after the seizure onset were normalized to have a maximum or minimum of 1 or -1 and fed into the trained DeepSIF model for source estimation. The power spectrum of 10 seconds before and after the ictal onset were calculated for each EEG channel and the major frequency for each seizure was manually identified by comparing the averaged EEG power-spectrum between pre- and post-seizure onset. The DeepSIF output was filtered at the major frequency with a passband of at most 5 Hz. The energy at each cortical segment was calculated over the three seconds and used as the spatial reconstruction of the seizure activity for evaluation. For interictal spikes analysis, 1 second of signal around the peak of the spikes were extracted and averaged for each patient and scaled by the maximum of the absolute value. The output source reconstruction was averaged for a 100 ms window around the peak of the spike. A threshold was determined using the Otsu’s thresholding technique [55] for ictal and interictal imaging results.

The spatial sensitivity, specificity and spatial dispersion (SD) were calculated with respect to the resection region. The harmonic and geometric mean of the sensitivity and specificity were calculated as 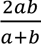 and 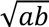 for *a* and *b* representing sensitivity and specificity. Spatial dispersion is defined as the weighted mean of the distance of each reconstructed region to the resection area. 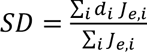, where *d_i_* is the minimum distance to the resection region for reconstructed region *i*, and *J_e_* is the estimated source map. The reconstructed waveforms in all reconstructed regions were averaged and the Pearson correlation between the averaged reconstructed waveform and the EEG channel with the maximum energy was calculated. When comparing to the SOZ electrodes, two LEs are calculated. The first LE is defined as the average distance of iEEG defined SOZ electrodes to the closest estimated source regions. The second LE is defined as the average distance of each estimated source region to the closest SOZ electrode. The SOZ LE is defined as the average value of these two LEs.

The imaging results from three conventional methods were calculated for ictal signals: time domain standardized low resolution brain electromagnetic tomography (sLORETA) [61], frequency domain imaging with sLORETA (FDI) [18], and time domain linearly constrained minimum variance (LCMV) beamformer [62]. The denoised EEG signal was filtered at its major frequency with a passband of at most 5 Hz and used as the input for the time-domain methods. After filtering, the EEG channel with the maximum energy was used to detect the local maxima. Local maxima with magnitude larger than 0.5 of the global maxima were assumed to be the peak of the ictal oscillations. The ESI results were averaged at the peak of the ictal oscillations to calculate the reconstructed spatial map [24]. The Fourier Transforms of the ictal EEG signal were calculated for each channel for the time domain method. The real and imagery part of the frequency domain signal at the major ictal frequency can be treated as two scalp maps and ESI can be performed for these two maps. The real and imaginary ESI results can be combined by calculating the magnitude of the complex number. The data analysis results for sLORETA and LCMV were calculated using MNE-Python (version 0.22.0) [63]. The Otsu’s method was used to determine the extent of the source imaging solution. One-sided Wilcoxon signed rank test and rank sum test were used for the statistical tests.

## Data Availability

All data produced in the present work are contained in the manuscript and supplementary information.

## Acknowledgement

This work was supported in part by National Institutes of Health grants NS127849 and NS096761, and by a gift from the Pittsburgh Health Data Alliance. The work used the Extreme Science and Engineering Discovery Environment (XSEDE), which was supported by National Science Foundation grant number ACI1548562. Specifically, it used the Bridges-2 system, which is supported by NSF award number ACI-1928147, at the Pittsburgh Supercomputing Center (PSC).

The authors are grateful to Dr. Ben Brinkmann and Ms. Cindy Nelson for assistance in clinical data collection and preparation, as well as Dr. Shuai Ye and Mr. Xiyuan Jiang for assistance in preparation of data.

## Contributions

R.S. and B.H. contributed to the conception of the project, developing analysis algorithms and performing data analysis. G.A.W. and B.J. contributed to the data collection and preparation. R.S., A.S., B.J., G.A.W., and B.H. contributed to the conduction of research. B.H. contributed to the supervision of the project. R.S. and B.H. contributed to the initial draft of the manuscript. All authors contributed to the revision of the manuscript.

