## Supplementary Information for "Spatiotemporal Rhythmic Seizure Sources Can be Imaged by means of Biophysically Constrained Deep Neural Networks"

### Supplementary Information
# of

### Spatiotemporal Rhythmic Seizure Sources Can be Imaged by means of Biophysically Constrained Deep Neural Networks


Rui Sun[1], Abbas Sohrabpour[1], Boney Joseph[2], Gregory Worrell[2], Bin He[1,3,*]

1. Department of Biomedical Engineering, Carnegie Mellon University, Pittsburgh, PA
2. Department of Neurology, Mayo Clinic, Rochester, MN
3. Neuroscience Institute, Carnegie Mellon University, Pittsburgh, PA

* Correspondence:

Bin He, PhD
Carnegie Mellon University
Department of Biomedical Engineering
5000 Forbes Avenue, Pittsburgh, PA 15213, USA


**Table S1**

#### Patient information

| Pt. # | Gender | Surgical Resection | Intracranial EEG | Follow-up | Outcome | Surgery age range |
|---|---|---|---|---|---|---|
| 1 | F | Right anterior temporal lobectomy and amygdalohippocampectomy | Right temporal grids and strips and depth electrodes on mesial side | 1 year | ILAE-1 | 31-35 |
| 2 | M | Right anterior temporal lobectomy and hippocampectomy | Bilateral depth electrodes into hippocampi | 3 years | ILAE-1 | 26-30 |
| 3 | F | Left anterior temporal lobectomy and amygdalohippocampectomy | Left frontal and temporal grids and strips and depth electrodes on mesial side | 3 years | ILAE-1 | 26-30 |
| 4 | F | Left anterior temporal lobectomy and amygdalohippocampectomy | Left temporal strips and mesial depth electrodes | 32 months | ILAE-1 | 21-25 |
| 5 | F | Left anterior frontal lobe excision | N.A. | 30 months | ILAE-1 | 21-25 |
| 6 | F | Right anterior temporal lobectomy and amygdalohippocampectomy | Right temporal grids and strips and depth electrodes on mesial side | 1 year | ILAE-1 | 21-25 |
| 7 | F | Left anterior temporal lobectomy and amygdalohippocampectomy | Left frontal and temporal grids and strips and mesial depth electrodes | 1 year | ILAE-2 | N.A. |
| 8 | F | Right temporal lobectomy and amygdalohippocampectomy | N.A. | 2 years | ILAE-2 | 46-50 |
| 9 | F | Right temporal lobectomy and amygdalohippocampectomy | Right temporal grids and strips and depth electrodes on mesial side | 1 year | ILAE-1 | 16-20 |
| 10 | F | Left temporal lobectomy and amygdalohippocampecomy | N.A. | 32 months | ILAE-1 | 56-60 |
| 11 | F | Left temporal lobectomy and amygdalohippocampecomy | Right temporal grids and strips and depth electrodes on mesial side | 1 year | ILAE-1 | 51-55 |
| 12 | F | Right temporal lobectomy and amygdalohippocampectomy | Right temporal grids and strips and depth electrodes on mesial side | 1 year | ILAE-1 | 21-25 |
| 13 | F | Extended right temporal lobectomy with amygdalohippocampectomy | Multiple right lateral temporal and sub-temporal grids and strips of electrodes | 15 months | ILAE-1 | 36-40 |

| | | | | | | |
|---|---|---|---|---|---|---|
| 14 | M | Right temporal lobectomy and amygdalohippocampectomy | Right temporal grids and strips and depth electrodes on mesial side | 1.5 years | ILAE-1 | 46-50 |
| 15 | M | Right anterior temporal lobectomy and amygdalohippocampectomy | Right temporal grids and strips and depth electrodes on mesial side | 1 year | ILAE-1 | 51-55 |
| 16 | F | Left parietal focal cortical resection | Right temporal grids and strips and depth electrodes on mesial side | 1 year | ILAE-1 | 21-25 |
| 17 | M | Modified left frontal lobectomy | Left frontal strips | 19 months | ILAE-1 | 21-25 |
| 18 | M | Right temporal lesion resection | N.A. | 19 months | ILAE-1 | 46-50 |
| 19 | F | Right frontal cortical resection | Right frontal, orbital frontal, and bilateral interhemispheric grid/stripes/depth electrodes | 14 months | ILAE-1 | 11-15 |
| 20 | F | Left anterior temporal lobectomy and amygdalohippocampectomy | Left parietal, left temporal mesial, and lateral neocortical region depth, subdural grid, and strip electrodes | 16 months | ILAE-1 | 21-25 |
| 21 | M | Right mesial frontal orbital lobe excision | Bilateral temporal and frontal depth electrodes | 15 months | ILAE-1 | 21-25 |
| 22 | M | Right frontotemporal lobe resection | Right temporoparietal and right parietal grid, right temporal depth electrodes | 11 months | ILAE-5 | 21-25 |
| 23 | M | Right temporal lobectomy and amygdalohippocampectomy | Bitemporal depth electrodes | 8 months | ILAE-4 | 36-40 |
| 24 | F | Right temporal lobectomy and amygdalohippocampectomy | Right temporal strips and mesial temporal depth electrodes | 19 months | ILAE-3 | 46-50 |
| 25 | M | Right anterior temporal lobectomyand amygdalohippocampectomy | Right temporal strips and mesial temporal depth electrodes | 18 months | ILAE-4 | 26-30 |
| 26 | F | Left anterior temporal lobectomy and amygdalohippocampectomy | Left temporal grid and strips on the inferior temporal side and left frontal, depth electrodes on temporal and frontal | 12 months | ILAE-4 | 21-25 |
| 27 | F | Focal temporo-occipital cortical resection | Right fronto-temporal grids and strips and depth electrodes close to amygdala and hippocampus, and later right posterotemporo-parietal grids and strips | 12 months | ILAE-4 | 21-25 |

| | | | | | | |
|---|---|---|---|---|---|---|
| 28 | M | Right temporal lobectomy and amygdalohippocampectomy | Right temporal strips | 13 months | ILAE-4 | 26-30 |
| 29 | F | Left temporal lobectomy and amygdalohippocampectomy | left temporal strips and mesial depth electrodes | 15 months | ILAE-4 | 56-60 |
| 30 | F | Left anterior temporal lobectomy and amygdalohippocampectomy | Left temporal strips and electrodes | 12 months | ILAE-3 | N.A. |
| 31 | M | Lateral temporal extending into parietal cortex resection | Left subdural grids, strips, and depth electrodes | 14 months | ILAE-5 | 31-35 |
| 32 | M | Posterior superior margin of the sylvian fissure resection | Right subdural grids, strips, and depth electrodes | 15 months | ILAE-5 | 36-40 |
| 33 | M | Left anterior temporal lobectomy, amygdalohippocampectomy, and left posterior temporo-occipital corticectomy | Left temporal subdural grids, strip electrodes, and left depth electrodes | 15 months | ILAE-3 | 46-50 |

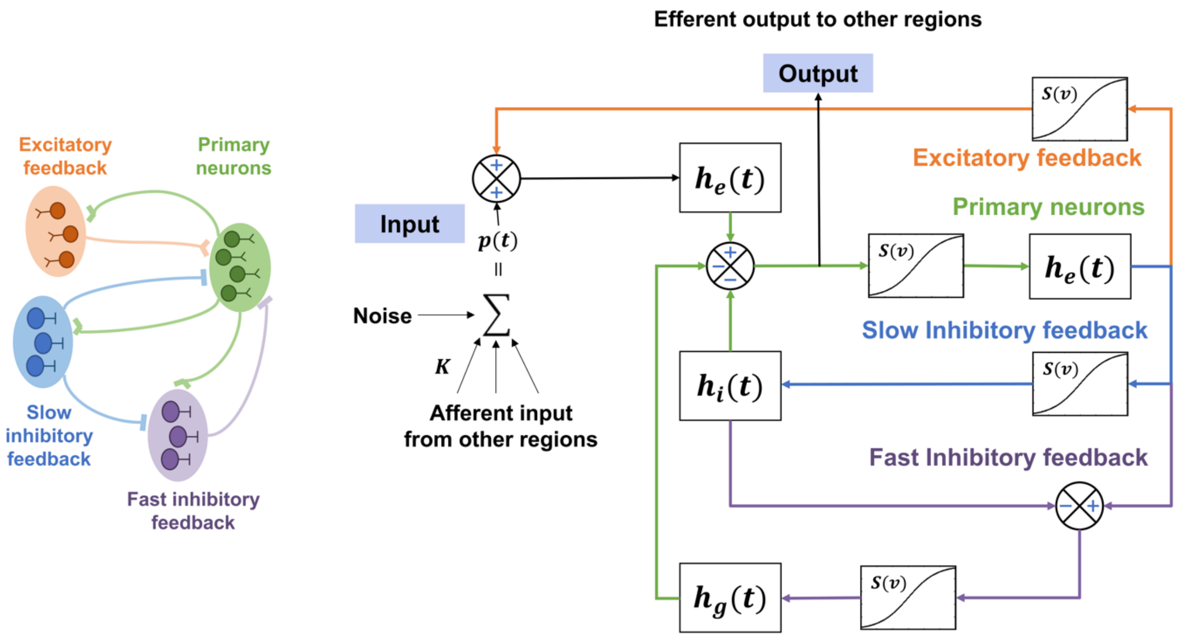


***Figure S1***. *A single-element modified Jansen-Rit model simulates four neuronal sub-populations: the primary neurons (i.e., pyramidal cells), as well as excitatory, slow inhibitory and fast inhibitory interneurons. The influence from other neural ensembles is modeled as an excitatory input p(t) that represents the average pulse density, i.e., average firing rate, of afferent action potentials. The behavior of each subpopulation is modeled using linear functions* $h(t)$ *that describe how presynaptic information, from excitatory and inhibitory populations, is transformed into postsynaptic information, and nonlinear function S(v) that models the change from average membrane potential of the subpopulation to average pulse density.*

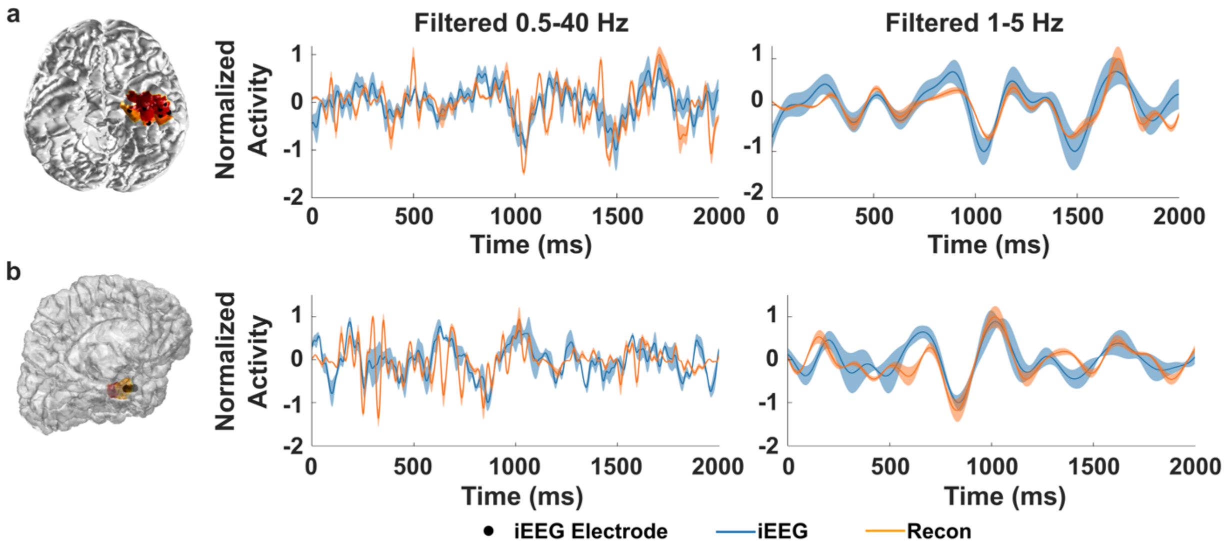


***Figure S2***. *Comparing estimated time-courses from EEG to intracranial EEG recordings in 2 patients. The reconstructed time-courses near the SOZ electrodes from DeepSIF were averaged and compared to the averaged iEEG traces. The major ictal frequency for these patients is around 3 Hz.* ***a,*** *The linear correlation between the iEEG and reconstruction is 0.5 for wide-band signal, and 0.84 for narrow-band signal filter around the major frequency.* ***b,*** *The linear correlation between the iEEG and reconstruction is 0.4 for wide-band signal, and 0.77 for narrow-band signal filter around the major frequency.*

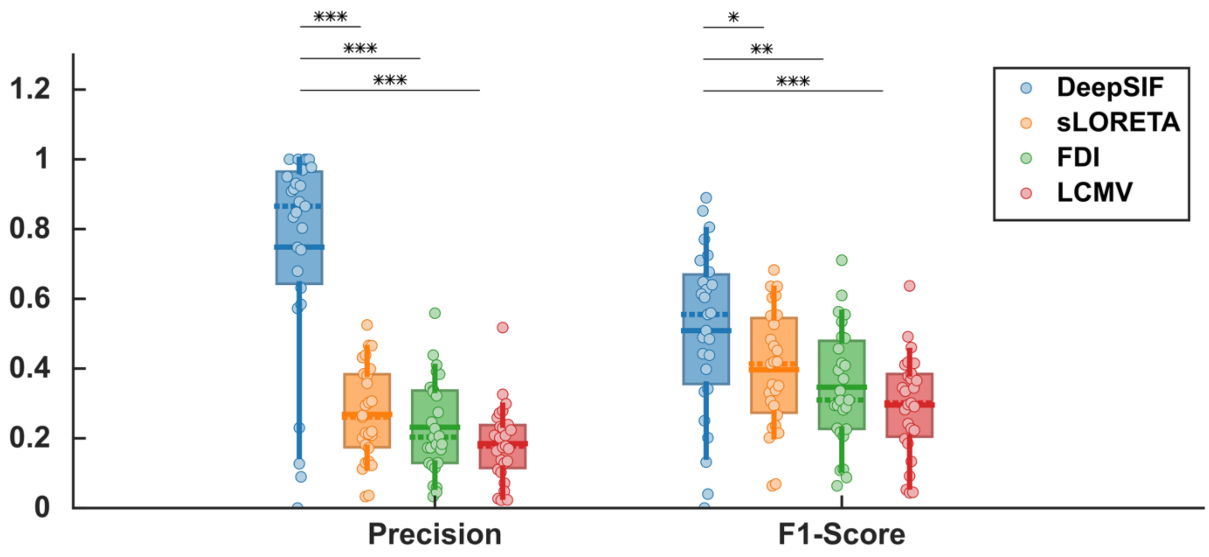


***Figure S3***. *Precision and F1-Score for different ictal source imaging methods. Paired one-sided Wilcoxon signed rank test was used with statistical significance cutoffs of (*P<0.05, **P<0.01, ***P<0.001).*

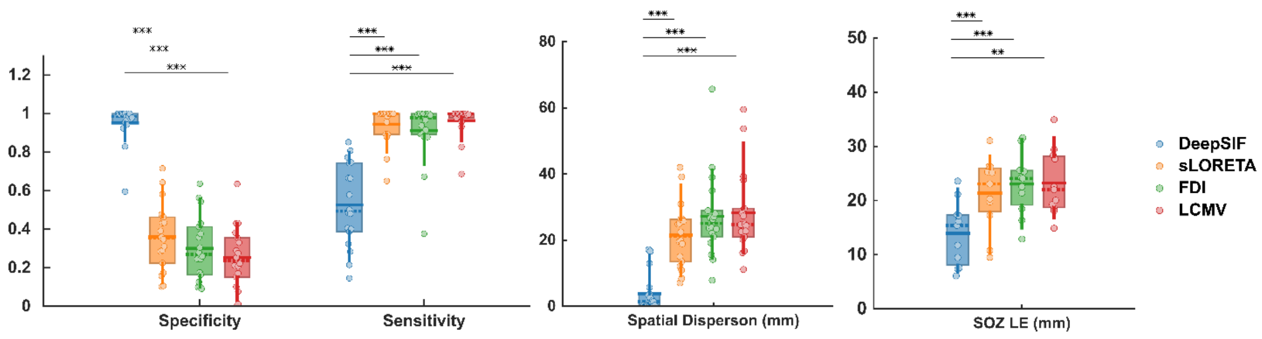


***Figure S4****. Ictal source imaging results for different ictal imaging methods in the seizure free group. Paired one-sided Wilcoxon signed rank test was used with statistical significance cutoffs of (*P<0.05, **P<0.01, ***P<0.001).*
